# MMSE-CDR-SB residual as an exploratory indicator of deviation from an Alzheimer’s disease-typical cognitive-functional pattern

**DOI:** 10.64898/2026.04.28.26351663

**Authors:** Ambre Mounié, Kenichiro Sato, Saki Nakashima, Masanori Kurihara, Ryoko Ihara, Yoshiki Niimi, Atsushi Iwata, Takeshi Iwatsubo

## Abstract

**INTRODUCTION:** We tested whether the MMSE-CDR-SB residual, defined as observed minus expected CDR-SB from an Alzheimer’s disease-oriented reference equation, reflects deviation from an AD-typical cognitive-functional pattern.

**METHODS:** Using NACC data, we analyzed an autopsy cohort (n=1,981) and a separate clinical diagnosis cohort (n=3,184). Associations were examined using multivariable logistic regression adjusted for MMSE band, age, sex, and education.

**RESULTS:** Lower residual values were associated with lower odds of amyloid and AD-type tau pathology. Positive residual values showed a directional but nonspecific association with selected non-AD co-pathologies, including TDP-43 and vascular burden. In the clinical cohort, positive residual values were enriched in PSP and FTLD-other, whereas AD cases clustered near the reference pattern.

**DISCUSSION:** The residual appears to summarize deviation from an AD-typical cognitive-functional pattern rather than indicate one specific pathology, and may serve as an exploratory triage-level signal of a less AD-typical presentation.

**Highlights:**

- MMSE-CDR-SB residual summarizes deviation from an AD-typical pattern.
- Lower residuals were associated with lower odds of amyloid and AD-type tau.
- Positive residuals were clinically enriched in PSP and FTLD-other.
- Residual has potential as a triage-level signal of a less AD-typical presentation.

**Research in Context:** *Systematic review:* We reviewed PubMed and reference lists for studies on MMSE-CDR-SB crosswalks, mixed dementia pathology, and syndrome-specific differences in cognitive-functional severity. Prior work mainly mapped scores across instruments and did not test whether deviation from an AD-oriented MMSE-CDR-SB reference relationship was associated with neuropathological or clinical heterogeneity.

*Interpretation:* In NACC, the MMSE-CDR-SB residual behaved as a summary of deviation from an AD-typical cognitive-functional pattern rather than as a specific pathology marker. Negative residuals were associated with lower odds of core AD pathology, whereas positive residuals were enriched in PSP and FTLD-other and showed only directional, nonspecific signals for selected non-AD co-pathologies.

*Future directions:* Prospective validation should test generalizability, refine practical thresholds, and determine whether this simple bedside metric improves etiologic triage for mixed pathology and non-AD syndromic features, especially where biomarker access is limited.

## Introduction

Dementia affects more than 55 million people worldwide and is a major cause of disability and dependency in older adults, with its prevalence expected to almost triple by 2050 [Nichols2022(PMID: 34998485);Mostert2025]. In clinical practice and therapeutic trials, cognitive and functional status are commonly assessed with two standardized tools: the Mini-Mental State Examination (MMSE), a brief screening measure of global cognitive performance [Folstein1975], and the Clinical Dementia Rating Sum of Boxes (CDR-SB), a composite measure of dementia severity integrating cognitive and functional impairment [Hughes1982; Morris1993(PMID: 8232972); O’Bryant2010]. These measures are also important for patient selection in Alzheimer’s disease (AD) disease-modifying therapies [Belder2023(PMID: 37463598);Cummings2023], including recent anti-amyloid treatments such as lecanemab and donanemab [Hartz2025].

In AD, the most common cause of dementia [Jalbert2008], MMSE and CDR-SB are generally correlated across disease stages. Brogaard et al. proposed a linear equation in the National Alzheimer’s Coordinating Center (NACC) dataset to estimate expected CDR-SB from MMSE in an AD-oriented framework: CDR-SB = −0.68 × MMSE + 20.2 [Brogaard2025]. Earlier studies also showed that common dementia scales such as MMSE, CDR-SB, and ADAS-Cog can be statistically linked across overlapping severity ranges [Balsis2015]. The Brogaard equation provides an expected CDR-SB value for a given MMSE score. Therefore, it can be used to quantify deviation from the AD-oriented reference relationship. In this study, we defined this deviation as the residual, calculated as the difference between observed and predicted CDR-SB. A positive residual means that the observed CDR-SB is higher than expected for a given MMSE score, suggesting greater overall cognitive-functional severity than predicted from cognition alone. A negative residual means that the observed CDR-SB is lower than expected, suggesting relatively lower overall clinical severity.

This residual may be clinically informative because different pathological processes may affect cognition and everyday function differently [Kapasi2017(PMID: 28488154)]. This idea is supported by studies showing that different dementia syndromes can show different levels of clinical or functional severity at similar levels of cognitive impairment. For example, FTLD has been reported to show greater clinical severity than AD after matching for MMSE and disease duration [Mioshi2017;Brown2011]. Therefore, deviation from the AD-oriented MMSE-CDR-SB reference relationship may serve as a practical summary of an atypical cognitive-functional presentation or possible mixed pathology, without implying that it provides information independent of MMSE and CDR-SB themselves. In this sense, the residual should be viewed not as a marker of one specific pathology, but as a simple summary of deviation from an AD-typical cognitive-functional pattern.

This question is clinically important because AD often co-occurs with other pathologies [Robinson2021; Kapasi2017]. In older adults, dementia is often a multi-morbid brain condition rather than the result of a single disease [DeReuck2016; Kapasi2017]. Frequent co-pathologies accompanying AD neuropathological change include Lewy body pathology, vascular pathology, cerebral amyloid angiopathy, and limbic-predominant age-related TDP-43 encephalopathy neuropathological change [Tosun2024]. These co-pathologies are clinically relevant because they contribute to cognitive decline and may reduce the measurable effect of therapies targeting only one pathological substrate [Tosun2024]. However, biomarkers for non-AD neuropathological changes are still limited in routine clinical practice [Tosun2024; Vos2024(PMID: 38724589)].

In routine practice, clinicians often encounter patients whose MMSE score and overall clinical severity do not fit a typical AD pattern, while access to biomarkers for non-AD co-pathologies remains limited. In this setting, a simple bedside measure is unlikely to establish etiology by itself. However, it may still be useful as an exploratory triage-level signal to identify presentations that appear less AD-typical and may merit closer etiologic evaluation. Prospectively, one possible use case would be to treat marked positive residuals as a prompt for closer non-AD syndromic or mixed-pathology evaluation, rather than as a diagnostic label.

Against this background, we used the AD-oriented MMSE–CDR-SB relationship as an interpretive benchmark and examined whether deviation from this benchmark differs across neuropathological and clinical subgroups. We did not treat the residual as an independent biological marker. Rather, we evaluated whether it may serve as a practical summary of a less AD-typical cognitive-functional presentation. Using the NACC dataset, we had two aims: first, to examine the association between the residual and neuropathological findings in an autopsy cohort; and second, to assess its relationship with clinician-assigned etiologic diagnoses in a separate clinical cohort.

## Methods

### Study design and participants

This study used data from the National Alzheimer’s Coordinating Center (NACC) Uniform Data Set (UDS), a multicenter longitudinal observational cohort that collects standardized clinical, cognitive, and neuropathological data from Alzheimer’s Disease Research Centers across the United States [Beekly2007]. We included visit data collected between 2005 and 2023. To reduce potential confounding by major non-neurodegenerative conditions that may affect cognition or function, participants were excluded if they had a history of stroke, Parkinson’s disease, alcohol abuse, schizophrenia, depressive disorder of less than two years’ duration, bipolar disorder, or a known autosomal dominant AD mutation.

For the cross-sectional analyses, we defined 2 analytic subcohorts according to data availability and study aim. For neuropathological analyses, we selected the clinical visit closest to death and retained only visits occurring within 24 months before death, in order to maximize the temporal proximity between ante-mortem clinical assessment and post-mortem findings. For clinical diagnosis analyses, we selected the visit closest to death with a clinician-reported etiologic diagnosis. We did not impose an upper limit on the interval to death in the main clinical analysis, primarily to preserve sample size. Because this design differs from the neuropathological cohort, we additionally evaluated the distribution of the visit-to-death interval in the clinical cohort and performed a sensitivity analysis restricting the clinical cohort to visits within 24 months before death.

Because the MMSE-CDR-SB reference equation proposed by Brogaard et al. was reported to perform less well in more severe stages, the main analyses were restricted to visits with MMSE scores between 16 and 30 [Brogaard2025]. After this restriction, the final analytic samples consisted of 1,981 participants in the neuropathological cohort and 3,184 participants in the clinical cohort (Figure S1). Cognitively normal individuals within this MMSE-eligible range were retained in both cohorts to preserve the observed cognitive-functional spectrum. These single-visit cohort definitions were used for the primary cross-sectional analyses.

### Discrepancy Index

For each eligible visit, we calculated the difference between observed CDR-SB and the value predicted from MMSE by the Brogaard equation (CDR-SB = 20.2 − 0.68 × MMSE) [Brogaard2025]. This value was used as the discrepancy index (residual), defined as: residual = observed CDRSB − (20.2 − 0.68 × MMSE).

A positive residual indicates that the observed CDR-SB is higher than expected for a given MMSE score, corresponding to greater overall cognitive-functional severity than predicted from cognition alone. A negative residual indicates that the observed CDR-SB is lower than expected, corresponding to less overall clinical severity than expected.

For descriptive, stratified, and graphical analyses, the discrepancy index was categorized a priori into 4 ordinal bins (≤−4, −4 to 0, 0 to +4, and ≥+4). These categories were chosen for interpretability and were not based on empirical cutoffs. In addition, MMSE was categorized into 3 bands (16–21, 22–27, and 28–30) for stratified analyses.

### Neuropathological and Clinical variables

Neuropathological variables were defined from NACC autopsy form items using predefined study definitions informed by the NACC Neuropathology Coding Guidebook and contemporary consensus frameworks. The main neuropathological variables included amyloid pathology (A+) [Montine2012], AD-type tau pathology (T+) [Braak1991], non-AD tau pathology [Kovacs2016], neurodegeneration (N+) [Jack2018], Lewy body pathology [McKeith2017], TDP-43 pathology [Nelson2019], hippocampal sclerosis [Nelson2011], cerebral amyloid angiopathy [vanVeluw2020], and high vascular burden [Skrobot2016]. High vascular burden was defined as a study-specific composite across prespecified vascular lesion variables. The main variable TDP-43 should not be interpreted as a broad age-related TDP-43 measure. Given its derivation from a single composite item without regional specificity, it predominantly reflects FTLD-type TDP-43 pathology in this dataset. Findings related to this variable should therefore be read as a directional signal rather than a robust prevalence estimate. Detailed coding rules and source variables are provided in Table S1.

For the clinical cohort, presumptive etiologic diagnoses were derived from the clinician-reported primary etiologic diagnosis variable (*NACCETPR*) and converted into one-vs-rest binary flags for Alzheimer’s disease (AD), Lewy body disease (LBD), vascular dementia (VaD), FTLD-other, progressive supranuclear palsy (PSP), corticobasal degeneration (CBD), other neurological disorders, and cognitively normal status. These variables reflect NACC-recorded etiologic diagnoses rather than diagnoses re-adjudicated within the present study. Detailed coding is provided in Table S1.

### Statistical Analysis

All statistical analyses were conducted in R version 4.5.1. Analyses were performed separately in the neuropathological cohort and the clinical cohort using complete-case data for the variables required by each model. Two-sided p values < 0.05 were considered statistically significant. No formal multiplicity adjustment was applied because the analyses were exploratory and hypothesis-generating. Because both MMSE and residual were categorized for interpretability rather than empirical optimization, these analyses were intended primarily to describe pattern-level associations rather than to establish decision thresholds for individual-level use.

As descriptive analyses, we plotted subgroup-specific smoothed MMSE–CDR-SB trajectories with the Brogaard AD reference line overlaid (Figures 1 and 2) and examined prevalence across MMSE-band × residual-bin strata. Adjusted prevalence was estimated using logistic-regression standardization, with age at visit, sex, and years of education as covariates; crude proportions were reported when the adjusted model did not converge or analyzable sample size was too small. The primary cross-sectional analyses used separate multivariable logistic regression models for each binary neuropathological and clinical outcome, with MMSE band and residual bin as the main predictors and age at visit, sex, and years of education as covariates. The reference categories were the 0 to +4 residual bin and the 16–21 MMSE band. Odds ratios, 95% confidence intervals, and p values were estimated using cluster-robust standard errors clustered by participant (NACCID). Table 2 reports the residual-bin coefficients, and full model outputs are provided in Table S2.

**Figure 1:**
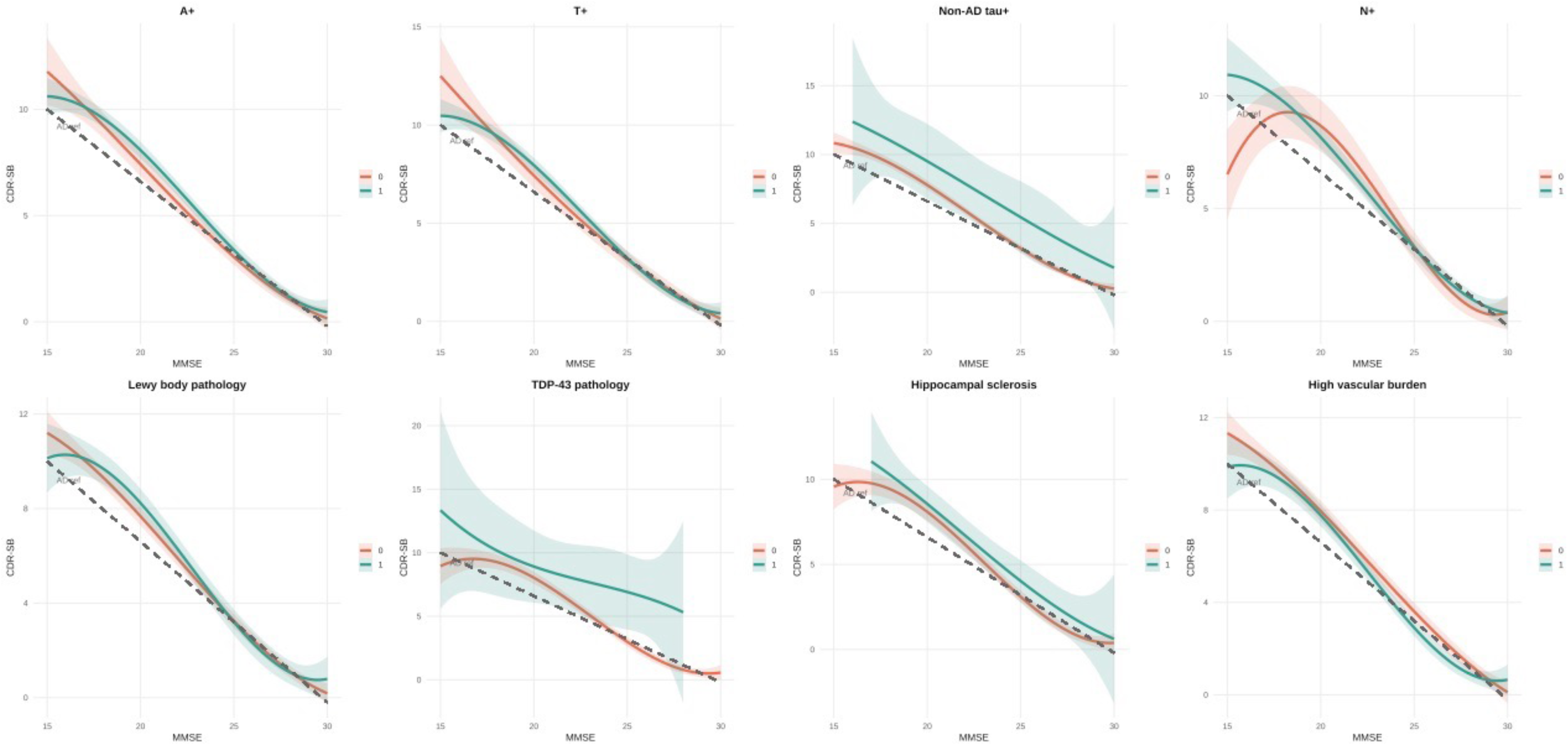
MMSE–CDR-SB relationship across neuropathological groups. Relationship between MMSE and CDR-SB across neuropathological categories (MMSE 15–30). Each panel shows smoothed regression curves (spline, 95% CI) for positive (1, green) vs. negative (0, red) groups. The reference trajectory for Alzheimer’s disease (AD), derived from Brogaard et al. (*CDR-SB = 20.2 − 0.68 × MMSE*), is overlaid (dashed line) for comparison. Shaded areas represent 95% confidence intervals around the smoothed trajectories. Only categories included in the formal GLM analysis (Table 2A) are shown; n ≥ 10 in both groups required. **Abbreviations**: A+, amyloid-positive; T+, tau-positive (AD pattern, Braak ≥ 3); Non-AD tau+, tau-positive (non-AD pattern); N+, neurodegeneration-positive; T class, tau− / AD-tau+ / nonAD-tau+.

**Figure 2.**
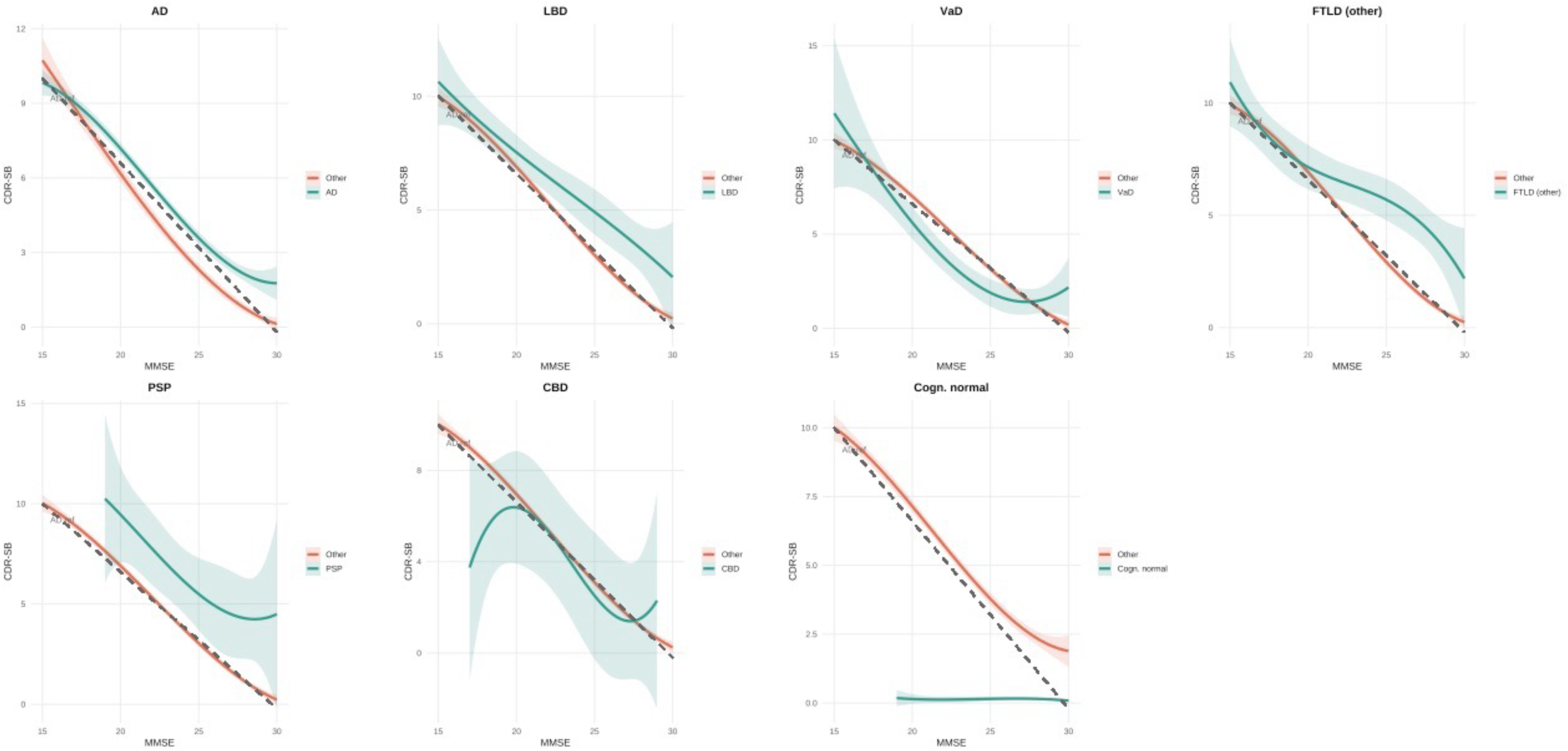
: MMSE–CDR-SB relationship across clinical diagnostic groups. Relationship between MMSE and CDR-SB across clinical etiologic diagnoses (MMSE 15–30). Each panel shows smoothed regression curves (spline, 95% CI) for the target diagnosis (1, green) vs. all others (0, red). Only categories with n ≥ 10 in both positive and negative groups are shown. The reference trajectory for Alzheimer’s disease (AD), derived from Brogaard et al. (*CDR-SB = 20.2 − 0.68 × MMSE*), is overlaid (dashed line) for comparison. Shaded bands represent 95% confidence intervals around the smoothed trajectories. **Abbreviations**: AD, Alzheimer’s disease; LBD, Lewy body dementia (clinical diagnosis; distinct from Lewy body pathology); VaD, vascular dementia; FTLD-other, frontotemporal lobar degeneration (other); PSP, progressive supranuclear palsy; CBD, corticobasal degeneration; Cogn. Normal, cognitively normal.

Sensitivity analyses for death/autopsy-related selection bias, analyses of co_AD × residual-bin interactions for selected pathologies, and exploratory longitudinal mixed-effects analyses are described in the Supplementary Methods; corresponding results are shown in Tables S3–S5 and Figure S5.

### Ethics

This study was approved by the University of Tokyo Graduate School of Medicine institutional ethics committee (ID: 2025264NI). Informed consent was not required because the study uses anonymized, publicly-available data only.

## Results

### Sample characteristics

The neuropathological cohort included 1,981 participants (mean age 83.1 ± 9.5 years; 46.7% female; mean education 15.6 ± 5.9 years), with a median visit-to-death interval of 10.2 months (IQR 5.9–14.8). The clinical cohort included 3,184 participants, with a median visit-to-death interval of 13.3 months (IQR 6.8–30.1). Mean residual values were close to zero in both cohorts (neuropathological cohort: 0.14 ± 2.62; clinical cohort: 0.09 ± 2.65), indicating that observed CDR-SB values were close on average to those predicted by the AD reference equation. Detailed participant characteristics are presented in Table 1.

**Table 1.** Basic Characteristics of Participants. Demographic, cognitive, and neuropathological characteristics are presented separately for the neuropathological cohort (n = 1,981) and the clinical cohort (n = 3,184). Time from visit to death is reported only for the neuropathological cohort, in which visits were restricted to within 24 months before death. The clinical cohort had no upper time limit on this interval. Percentages for neuropathological variables are based on variable-specific non-missing denominators, which vary substantially across variables due to differences in neuropathological form version availability. The total cohort n is provided in the column header for reference. The discrepancy index was computed as CDR-SB − (20.2 − 0.68 × MMSE), following Brogaard et al. Neuropathological variables were derived from standardized NACC autopsy data and are reported for the neuropathological cohort only. **Abbreviations:** SD, standard deviation; MMSE, Mini-Mental State Examination; CDR-SB, Clinical Dementia Rating Sum of Boxes; MCI, mild cognitive impairment; A+, amyloid positive; T+, AD-tau positive (Braak ≥ 3); N+, neurodegeneration positive; LBD, Lewy body disease; TDP-43, transactive response DNA-binding protein 43; HS, hippocampal sclerosis; CAA, cerebral amyloid angiopathy; PSP, progressive supranuclear palsy; CBD, corticobasal degeneration; FTLD, frontotemporal lobar degeneration. Discrepancy index, CDRSB−(20.2−0.68×MMSE). Reference category for discrepancy bin: 0 to +4.

| Characteristic | Neuropathological Cohort<br>(n = 1981) | Clinical Cohort<br>(n = 3184) |
| --- | --- | --- |
| **Demographics** |  |  |
| Age at visit, mean ± SD (years) | 83.1 ± 9.5 | 81.5 ± 10.2 |
| Female sex, % | 46.7% | 48.8% |
| Education, mean ± SD (years) | 15.6 ± 5.9 | 15.8 ± 7.5 |
| **Visit characteristics** |  |  |
| Time from visit to death, median [IQR] (months) | 10.2 [5.9-14.8] | 13.3 [6.8-30.1] |
| **Cognitive and Functional Scores** |  |  |
| MMSE, mean ± SD | 24.8 ± 4.2 | 24.4 ± 4.4 |
| CDR-SB, mean ± SD | 3.5 ± 4.1 | 3.7 ± 4 |
| **CDR Global Score, n (%)** |  |  |
| 0 | 647 (32.7%) | 938 (29.5%) |
| 0.5 | 667 (33.7%) | 1070 (33.6%) |
| 1 | 399 (20.1%) | 732 (23%) |
| 2 | 247 (12.5%) | 412 (12.9%) |
| 3 | 21 (1.1%) | 32 (1%) |
| **MMSE Band, n (%)** |  |  |
| 16–21 | 491 (24.8%) | 888 (27.9%) |
| 22–27 | 746 (37.7%) | 1200 (37.7%) |
| 28–30 | 744 (37.6%) | 1096 (34.4%) |
| **Residual Bin, n (%)** |  |  |
| $\leq -4$ | 67 (3.4%) | 132 (4.1%) |
| -4 to 0 | 976 (49.3%) | 1556 (48.9%) |
| 0 to +4 | 766 (38.7%) | 1226 (38.5%) |
| $\geq +4$ | 172 (8.7%) | 270 (8.5%) |
| **Neuropathological Variables, % positive (n)** |  |  |
| **Neuropathological Cohort only** |  |  |
| A+ (amyloid) | 54.1% (n= 1,023) | — |
| T+ (AD tau) | 62.9% (n= 1,010) | — |
| N+ (neurodegeneration) | 77.5% (n= 511) | — |
| Lewy body pathology | 22.5% (n= 1,020) | — |
| TDP-43 pathology | 4.2% (n= 428) | — |
| LATE-like (regional TDP) | 26.5% (n= 366) | — |
| LATE stage $\geq 2$ | 16.8% (n= 346) | — |
| Hippocampal sclerosis | 11.4% (n= 507) | — |
| High vascular burden | 46.4% (n= 1,023) | — |
| CAA (any) | 50.1% (n= 1,001) | — |
| **Presumptive Etiologic Diagnosis, n (%)** |  |  |
| **Clinical Cohort only** |  |  |
| Alzheimer's disease | — | 1525 (47.9%) |
| Lewy body disease | — | 119 (3.7%) |
| Vascular dementia | — | 109 (3.4%) |
| FTLD-other | — | 161 (5.1%) |
| PSP | — | 38 (1.2%) |
| CBD | — | 16 (0.5%) |
| Other neurological | — | 16 (0.5%) |
| Cognitively normal | — | 935 (29.4%) |

### Visual inspection of MMSE–CDR-SB trajectories across pathological and clinical groups

Descriptive MMSE–CDR-SB trajectories by subgroup are shown in Figures 1 and 2. Core AD-related pathological markers, including A+, T+, and N+, largely followed the Brogaard reference line. In contrast, TDP-43 pathology, and to a lesser extent hippocampal sclerosis, tended to lie above the reference line. Clinically, FTLD-other, PSP, and vascular dementia tended to lie above the reference line, whereas cognitively normal participants fell well below it. These descriptive patterns were evaluated formally in the regression analyses below.

### Association of the Residual Index with Neuropathological Markers

In cross-sectional regression analysis, the residual index showed significant overall associations with amyloid positivity, AD-tau positivity, high vascular burden, and TDP-43, whereas no significant overall association was observed for Lewy body pathology or neurodegeneration (Table 2A).

**Table 2.**
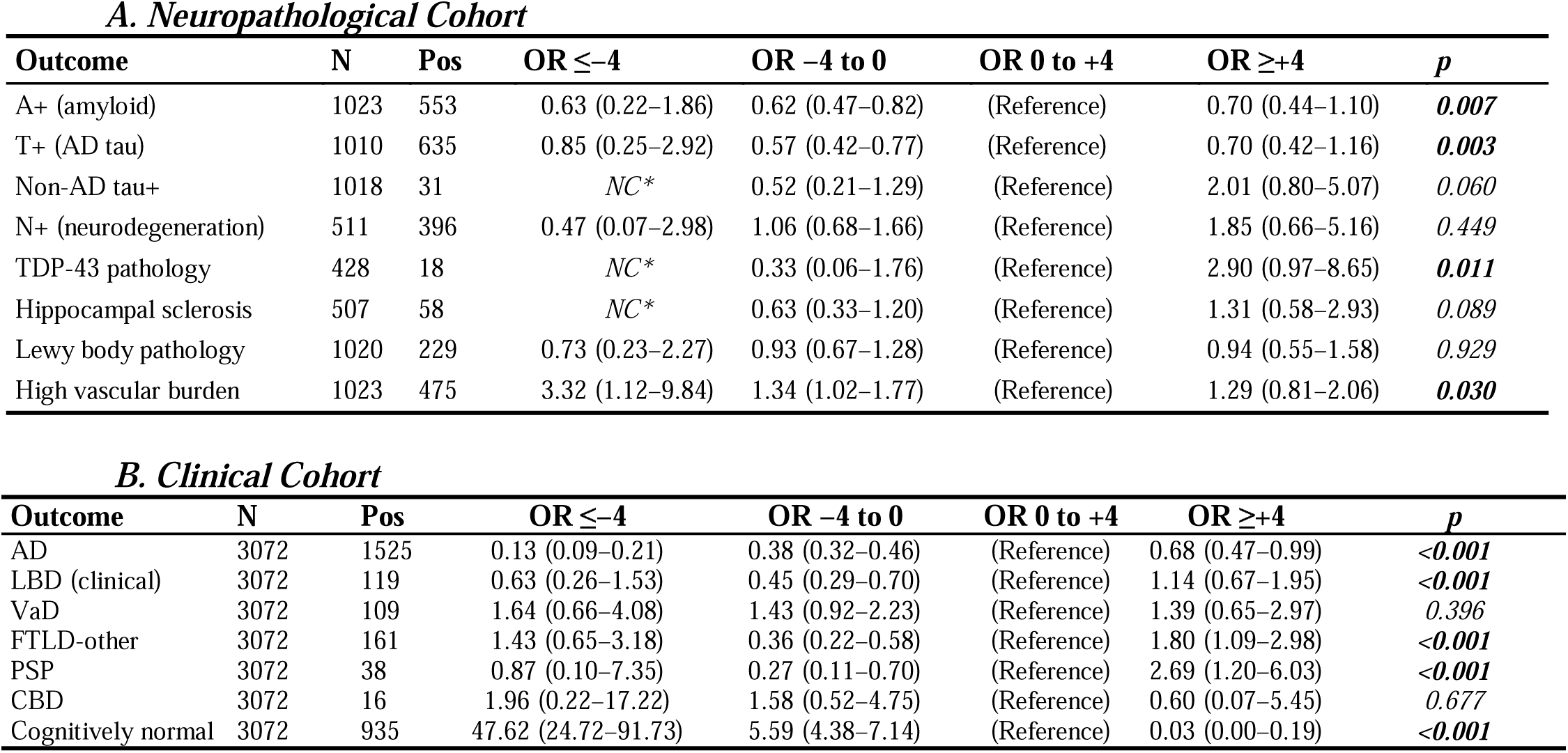
Adjusted Odds Ratio for Residual Bin. p (LRT): overall p-value from a likelihood ratio test comparing the full model with residual bin categories to a reduced model without residual bin, testing the joint association of all discrepancy categories with the outcome. Category-specific odds ratios and their p-values are reported in Table S2. **A** presents adjusted odds ratios (ORs) for the association between discrepancy index bins and the presence of neuropathological co-pathologies in the neuropathological cohort. CAA was not included in the primary regression analyses to maintain consistency with the pre-specified outcome set; it is described in Table 1 and Table S1. **B** presents the corresponding associations with presumptive clinical diagnoses in the clinical cohort. Odds ratios were estimated using logistic regression models with discrepancy bin as the primary predictor and the bin 0 to +4 as the reference category. All models were adjusted for age at visit, sex, and years of education. Odds ratios greater than 1 indicate higher odds of the outcome relative to the reference discrepancy bin, whereas values lower than 1 indicate lower odds. Other neurological disorders’ was excluded from the primary regression analyses owing to insufficient positive cases (n = 16). The neuropathological cohort includes participants with autopsy data and a clinical visit within 24 months prior to death. The clinical cohort includes participants with a clinician-reported etiologic diagnosis at their visit closest to death. *NC\** indicates “not calculable” due to zero cells / complete separation. Bold p-values indicate statistical significance (p < 0.05). **Abbreviations**: OR, odds ratio; CI, confidence interval; LRT, likelihood ratio test; PSP, progressive supranuclear palsy; CBD, corticobasal degeneration; FTLD, frontotemporal lobar degeneration.

| **Outcome** | **N** | **Pos** | **OR $\leq -4$** | **OR $-4$ to $0$** | **OR $0$ to $+4$** | **OR $\geq +4$** | ***p*** |
| --- | --- | --- | --- | --- | --- | --- | --- |
| A+ (amyloid) | 1023 | 553 | 0.63 (0.22–1.86) | 0.62 (0.47–0.82) | (Reference) | 0.70 (0.44–1.10) | ***0.007*** |
| T+ (AD tau) | 1010 | 635 | 0.85 (0.25–2.92) | 0.57 (0.42–0.77) | (Reference) | 0.70 (0.42–1.16) | ***0.003*** |
| Non-AD tau+ | 1018 | 31 | NC* | 0.52 (0.21–1.29) | (Reference) | 2.01 (0.80–5.07) | *0.060* |
| N+ (neurodegeneration) | 511 | 396 | 0.47 (0.07–2.98) | 1.06 (0.68–1.66) | (Reference) | 1.85 (0.66–5.16) | *0.449* |
| TDP-43 pathology | 428 | 18 | NC* | 0.33 (0.06–1.76) | (Reference) | 2.90 (0.97–8.65) | ***0.011*** |
| Hippocampal sclerosis | 507 | 58 | NC* | 0.63 (0.33–1.20) | (Reference) | 1.31 (0.58–2.93) | *0.089* |
| Lewy body pathology | 1020 | 229 | 0.73 (0.23–2.27) | 0.93 (0.67–1.28) | (Reference) | 0.94 (0.55–1.58) | *0.929* |
| High vascular burden | 1023 | 475 | 3.32 (1.12–9.84) | 1.34 (1.02–1.77) | (Reference) | 1.29 (0.81–2.06) | ***0.030*** |

***B. Clinical Cohort***
| **Outcome** | **N** | **Pos** | **OR $\leq -4$** | **OR $-4$ to $0$** | **OR $0$ to $+4$** | **OR $\geq +4$** | ***p*** |
| --- | --- | --- | --- | --- | --- | --- | --- |
| AD | 3072 | 1525 | 0.13 (0.09–0.21) | 0.38 (0.32–0.46) | (Reference) | 0.68 (0.47–0.99) | ***<0.001*** |
| LBD (clinical) | 3072 | 119 | 0.63 (0.26–1.53) | 0.45 (0.29–0.70) | (Reference) | 1.14 (0.67–1.95) | ***<0.001*** |
| VaD | 3072 | 109 | 1.64 (0.66–4.08) | 1.43 (0.92–2.23) | (Reference) | 1.39 (0.65–2.97) | *0.396* |
| FTLD-other | 3072 | 161 | 1.43 (0.65–3.18) | 0.36 (0.22–0.58) | (Reference) | 1.80 (1.09–2.98) | ***<0.001*** |
| PSP | 3072 | 38 | 0.87 (0.10–7.35) | 0.27 (0.11–0.70) | (Reference) | 2.69 (1.20–6.03) | ***<0.001*** |
| CBD | 3072 | 16 | 1.96 (0.22–17.22) | 1.58 (0.52–4.75) | (Reference) | 0.60 (0.07–5.45) | *0.677* |
| Cognitively normal | 3072 | 935 | 47.62 (24.72–91.73) | 5.59 (4.38–7.14) | (Reference) | 0.03 (0.00–0.19) | ***<0.001*** |

For the core AD pathological markers, participants with residuals below the reference value were less likely to harbor amyloid or AD-type tau pathology than those in the reference residual bin (Table S2), consistent with lower overall clinical severity than expected for a given level of cognitive impairment.

For TDP-43 pathology, the direction of association was consistent with a priori hypothesis. Participants in the highest residual bin (≥+4) had higher odds of TDP-43 positivity relative to the reference bin (OR = 2.90, 95% CI: 0.97–8.65, category-specific p = 0.057), although the overall likelihood ratio test for the residual bin term was significant (p = 0.057, Table 2A). Interpretation is limited by the small number of TDP-43-positive cases and sparse data in the lowest residual bin, in which no TDP-43-positive cases were observed (Figure 3). High vascular burden was also associated with the residual index, but the pattern was spread across multiple residual categories rather than confined to the positive extreme, suggesting a broader and less specific relationship. Complete regression results are provided in Table S2.

**Figure 3:**
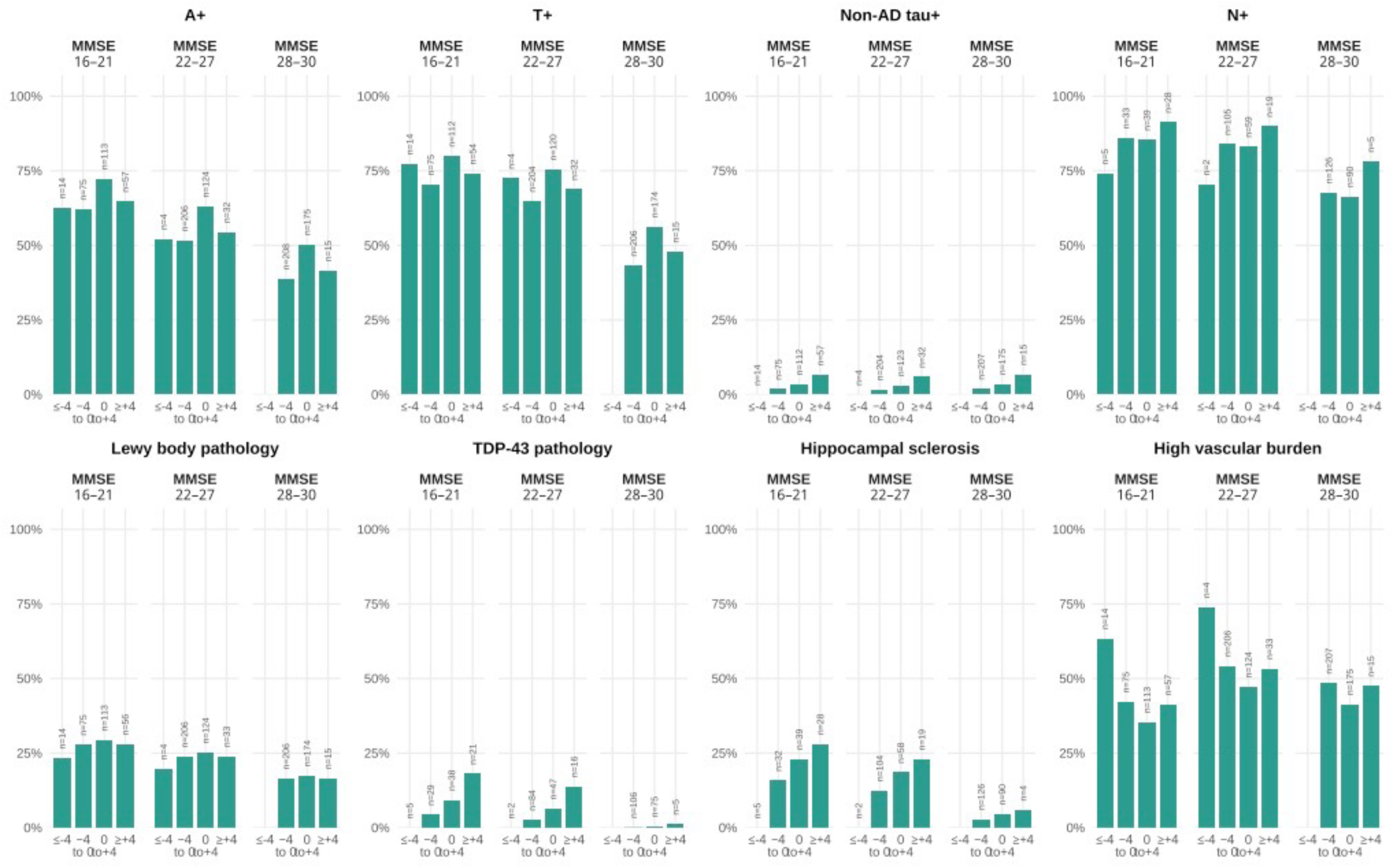
Prevalence of neuropathological co-pathologies across discrepancy bins, stratified by MMSE band. Bar plots display the age-, sex-, and education-adjusted proportion of participants positive for each neuropathological variable across categories of discrepancy between observed and expected CDR-SB given MMSE (discrepancy bins: ≤ −4, −4 to 0, 0 to +4, ≥ +4), stratified by MMSE band (16–21, 22–27, 28–30). Adjusted proportions were estimated using logistic standardization based on models including discrepancy bin, MMSE band, age at visit, sex, and years of education. Predicted probabilities were averaged across the observed covariate distribution within each MMSE band × discrepancy bin cell. The number of participants in each bin is displayed within each bar. Only variables with n ≥ 10 cases overall and used in the GLM are shown. Sample sizes (n) are indicated within each bar. Analyses were conducted in the neuropathological cohort (n = 1,981). **Abbreviations:** CDR-SB, Clinical Dementia Rating Scale Sum of Boxes; MMSE, Mini-Mental State Examination; A+, amyloid-positive (Thal phase ≥ 3 or CERAD score ≥ 2 or ADNC ≥ 2); T+, AD-tau positive (Braak stage ≥ 3); Non-AD tau+, tau-positive in a non-AD distribution; N+, neurodegeneration-positive.

**Figure 4:**
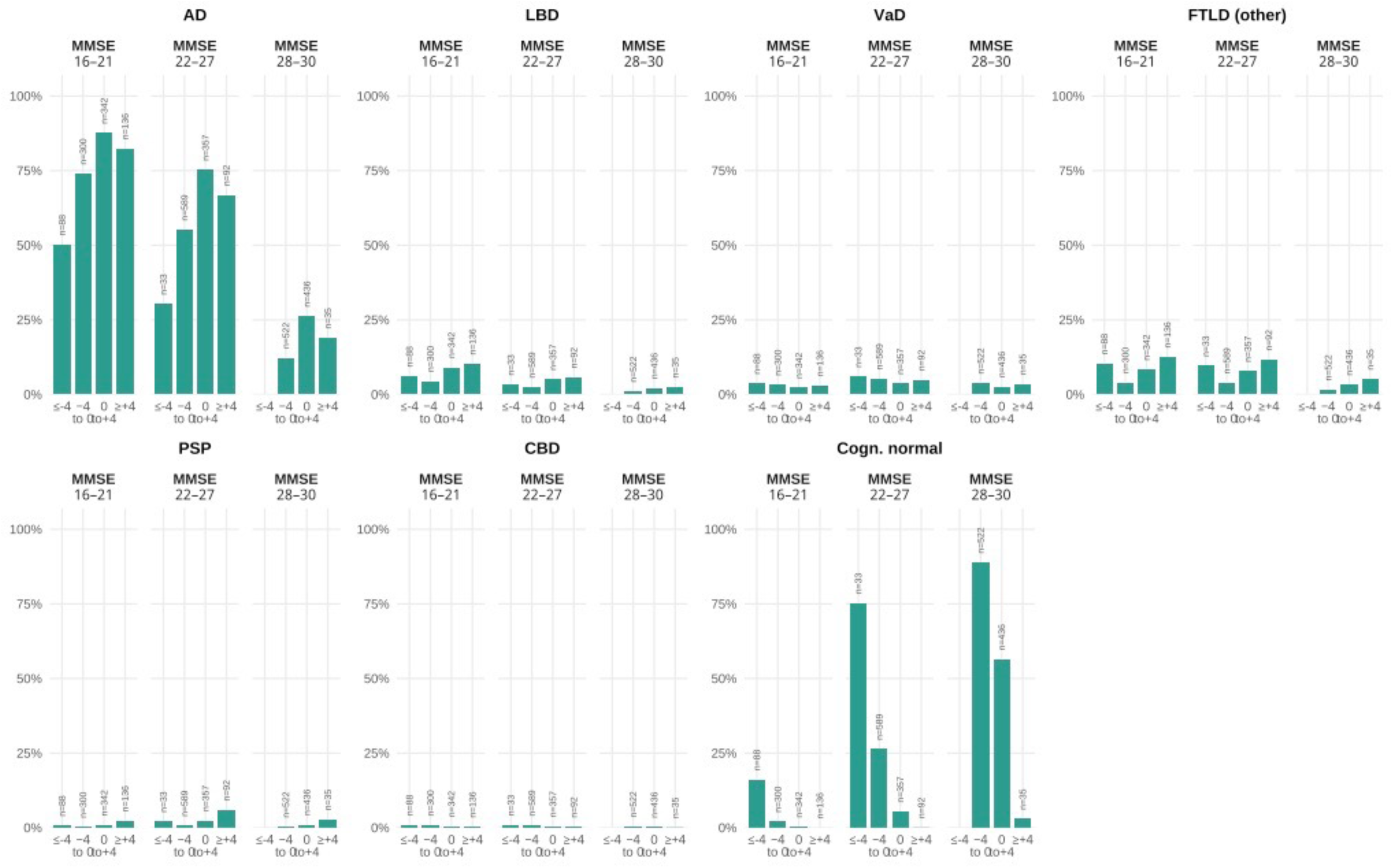
Prevalence of presumptive etiologic diagnoses across discrepancy bins, stratified by MMSE band. Bar plots display the age-, sex-, and education-adjusted proportion of participants with each clinician-reported etiologic diagnosis across categories of discrepancy between observed and expected CDR-SB given MMSE (discrepancy bins: ≤ −4, −4 to 0, 0 to +4, ≥ +4), stratified by MMSE band (16–21, 22–27, 28–30). Adjusted proportions were estimated using logistic standardization based on models including discrepancy bin, MMSE band, age at visit, sex, and years of education. Predicted probabilities were averaged across the observed covariate distribution within each MMSE band × discrepancy bin cell. Only diagnoses with n ≥ 10 cases overall are shown. Sample sizes (n) are indicated within each bar. Analyses were conducted in the clinical cohort (n = 3,184). **Abbreviations:** CDR-SB, Clinical Dementia Rating Scale Sum of Boxes; MMSE, Mini-Mental State Examination; AD, Alzheimer’s disease; LBD, Lewy body dementia (clinical diagnosis); VaD, vascular dementia; FTLD-other, frontotemporal lobar degeneration (other subtypes); PSP, progressive supranuclear palsy; CBD, corticobasal degeneration; Cogn. normal, cognitively normal.

### Association of the Residual Index with Clinician-Assigned Etiologic Diagnosis

In the clinical cohort, the residual index was significantly associated with several clinician-assigned etiologic diagnoses. The most clinically interpretable associations were observed for AD, PSP, and FTLD-other (Table 2B).

Cognitively normal status was concentrated in the lower residual range, with substantially higher odds in the negative residual bins. Conversely, AD diagnosis was less common away from the reference bin, including in the highest residual category (OR for ≥+4 bin vs reference, 0.68, 95% CI: 0.47–0.99, p = 0.043), consistent with clinically typical AD clustering near the AD-derived reference relationship.

PSP showed a positive residual pattern, with higher odds in the ≥+4 bin (OR = 2.69, 95% CI: 1.20–6.03, p = 0.016) and lower odds in the −4 to 0 bin (OR = 0.27, 95% CI: 0.11–0.70, p = 0.007). A similar pattern was observed for FTLD-other, with higher odds in the ≥+4 bin (OR = 1.80, 95% CI: 1.09–2.98, p = 0.021) and lower odds in the −4 to 0 bin (OR = 0.36, 95% CI: 0.22–0.58, p < 0.001). No clear association was observed for vascular dementia (overall p = 0.396). For LBD, the overall association reached significance, but this was mainly driven by lower odds in the negative residual range rather than enrichment in the positive range. CBD was not significantly associated with the residual index (overall p = 0.677). Overall, these findings suggest that the residual index captures clinical heterogeneity, with positive residuals enriched in selected non-AD syndromic diagnoses such as PSP and FTLD-other.

### Sensitivity Analysis for Death/Autopsy-Related Selection Bias

Death status was significantly associated with AD, FTLD-other, LBD, and cognitively normal status, and the death × residual-bin interaction was significant for AD, vascular dementia, FTLD-other, and cognitively normal status (Table S3). These findings indicate that death- and autopsy-related selection effects should be considered when interpreting the neuropathological analyses.

### Supplementary Analysis: Interaction with AD background

An exploratory analysis examining co_AD × residual-bin interactions for FTLD-like TDP, LATE-like, LATE stage ≥2, and high vascular burden showed no significant interaction for any outcome (all likelihood ratio test p > 0.05; Table S4, Figure S4). These analyses were limited by sparse data for some outcomes, especially TDP-related variables.

### Supplementary Analysis: Exploratory longitudinal analysis

In exploratory linear mixed-effects models, the residual index increased over time across groups. Steeper increases were observed in participants positive for amyloid, FTLD-like TDP-43, LATE-like, and LATE stage ≥2 pathologies, but not for high vascular burden (Table S5, Figure S5). These findings should be interpreted cautiously because pathological ascertainment was cross-sectional.

## Discussion

In this study, we examined whether deviation from an AD-oriented MMSE-CDR-SB reference relationship can be used as a practical summary of a less AD-typical cognitive-functional presentation. Our findings suggest that the residual is better understood as a summary of deviation from the AD-typical cognitive-functional pattern than as an independent pathology marker. Lower residual values were associated with lower odds of core AD pathology, whereas positive residual values were enriched in selected non-AD syndromic diagnoses, especially PSP and FTLD-other. In this sense, the residual may be most useful as a practical triage-level signal of a less AD-typical cognitive-functional presentation.

Among the neuropathological findings, vascular burden showed the strongest statistical association, but the pattern was not specific. Higher odds were seen in more than one residual category, so this finding does not suggest one clear biological direction. By contrast, TDP-43 pathology showed a more consistent directional pattern. Participants in the highest residual bin had higher odds of TDP-43 positivity, and no TDP-43-positive cases were seen in the lowest residual bin. This result should be interpreted carefully because the number of positive cases was small and missing data were common. Moreover, the TDP-43 variable used here is derived from a single composite NACC item and leans toward FTLD-type TDP-43 rather than reflecting the broader age-related TDP-43 construct associated with LATE. It should therefore not be generalized to age-related TDP-43 pathology. Still, the direction of the association is biologically plausible, because TDP-43 pathology often affects limbic structures and may increase memory-related and functional impairment more than would be expected from MMSE alone [Josephs2015; Meneses2021]. This pattern is consistent with the view that TDP-43-related pathology may create a dissociation between cognitive and functional severity that is captured by the deviation from the AD-typical MMSE-CDR-SB relationship. If TDP-43 adds limbic and functional burden to AD-related change, a positive residual may reflect this additional burden.

Most other neuropathological markers showed weak or inconsistent associations with the residual index. This is not surprising, because dementia in older adults is usually related to mixed and overlapping pathologies [Kapasi2017; Robinson2021; DeReuck2016]. Many lesions may contribute to clinical severity, but they do not always create a clear dissociation between cognition and function [Kapasi2017; Tosun2024]. Therefore, the residual index should not be expected to map directly onto every individual pathology. This is also consistent with the current view that dementia presentation in older adults is shaped by combinations of lesions rather than by one isolated process. For this reason, a simple residual measure is more likely to reflect overall heterogeneity of disease expression than a specific lesion-by-lesion correspondence.

The clinical cohort gave an important complementary result. In this analysis, AD cases tended to stay near the AD-derived reference line, while cognitively normal participants were concentrated in the negative residual range. In contrast, positive residual values were more common in non-AD syndromic diagnoses such as PSP and FTLD-other. These findings suggest that the residual index may capture atypical cognitive-functional profiles in clinical practice [Mioshi2017]. This interpretation is in line with earlier studies suggesting that functional or global clinical severity can differ across syndromes even at similar levels of cognitive impairment [Mioshi2017;Brown2011;Teng2010]. In this sense, the residual may be useful not because it identifies one diagnosis, but because it highlights patients whose clinical profile looks less AD-typical. However, there was still a large overlap between diagnostic groups. For this reason, the residual index alone is not enough for etiologic diagnosis at the individual level, and it should be considered only as an additional clinical signal [Devenney2017]. This limitation is not surprising, because the MMSE is a brief global screening tool and the reference equation itself was derived in an AD-oriented setting. Therefore, the residual should be interpreted as an additional clinical clue, not as a replacement for syndrome-based assessment or biomarker evaluation. Its main practical value may be to help prioritize which patients warrant closer etiologic workup when the presentation is less typical of clinically typical AD.

Clinically, the main use of this residual may be triage rather than diagnosis at the individual level. It should be noted that the residual is a mathematical combination of MMSE and CDR-SB and does not add information that is fully independent from these two measures. Its value lies in summarizing their joint deviation from an AD-oriented reference, not in capturing a distinct biological signal. This use should be understood as a screening-level application rather than a rule-based diagnostic threshold, because the residual categories were chosen for interpretability and were not empirically optimized for individual decision making. This may be particularly relevant in settings where biomarker access is limited and clinicians need a simple bedside cue to prioritize additional evaluation.

Several limitations should be noted. First, the neuropathological analyses were based on an autopsy-defined cohort and were therefore vulnerable to selection bias [Gauthreaux2024]. Our sensitivity analyses also suggested that death-related selection may affect how residual categories relate to diagnosis. Second, the apparently low frequency of TDP-43 positivity in this older AD-predominant cohort may reflect not only biology but also the variable definition, because the main TDP variable mainly reflects FTLD-type rather than age-related TDP-43, as well as substantial missingness, form-version dependence, and autopsy selection bias. Therefore, this finding should be interpreted as hypothesis-generating rather than as a robust prevalence estimate. Third, MMSE is a brief global screening test and does not capture domain-specific deficits in detail [Devenney2017]. This is particularly relevant for disorders such as LBD, in which visuospatial and neuropsychiatric features are prominent [McKeith2017], and vascular pathology, which often affects executive function more than memory [Skrobot2016]. Accordingly, the residual framework may be less sensitive for non-AD conditions requiring multidimensional assessment. In addition, NACC data may not fully capture functional or psychiatric comorbidities that could independently increase CDR-SB. Fourth, residual confounding remains possible, and the residual itself is a linear function of MMSE and CDR-SB. It therefore does not provide information independent of the original measures and should be viewed as a practical summary index rather than a separate biological construct [Elman2022]. Two mechanical issues also require caution: CDR-SB floor effects may partly explain the negative residual pattern in cognitively normal participants, and near MMSE ≈ 30 the linear equation can yield slightly negative predicted CDR-SB values. Finally, because the reference MMSE-CDR-SB equation was derived in an AD-oriented framework [Kovacs2016], the clustering of AD cases near the reference pattern is partly expected by construction and should not be overinterpreted as diagnostic specificity. In this study, the residual should be interpreted as deviation from an AD-typical benchmark rather than from a universal dementia trajectory [Kovacs2016; Kapasi2017].

In conclusion, the MMSE-CDR-SB residual may serve as a simple summary of deviation from an AD-typical cognitive-functional pattern. In this study, lower residual values were associated with lower odds of core AD pathology, whereas positive residual values were enriched in selected non-AD syndromic diagnoses such as PSP and FTLD-other. These findings support the residual as an exploratory triage-level signal rather than as a standalone diagnostic or prognostic marker. Future work should clarify its performance in time-restricted clinical cohorts and prospectively evaluate whether it adds practical value for etiologic triage.

## Acknowledgements

The NACC database is funded by NIA/NIH Grant U24 AG072122. NACC data are contributed by the NIA-funded ADRCs: P30 AG062429 (PI James Brewer, MD, PhD), P30 AG066468 (PI Oscar Lopez, MD), P30 AG062421 (PI Bradley Hyman, MD, PhD), P30 AG066509 (PI Thomas Grabowski, MD), P30 AG066514 (PI Mary Sano, PhD), P30 AG066530 (PI Helena Chui, MD), P30 AG066507 (PI Marilyn Albert, PhD), P30 AG066444 (PI David Holtzman, MD), P30 AG066518 (PI Lisa Silbert, MD, MCR), P30 AG066512 (PI Thomas Wisniewski, MD), P30 AG066462 (PI Scott Small, MD), P30 AG072979 (PI David Wolk, MD), P30 AG072972 (PI Charles DeCarli, MD), P30 AG072976 (PI Andrew Saykin, PsyD), P30 AG072975 (PI Julie A. Schneider, MD, MS), P30 AG072978 (PI Ann McKee, MD), P30 AG072977 (PI Robert Vassar, PhD), P30 AG066519 (PI Frank LaFerla, PhD), P30 AG062677 (PI Ronald Petersen, MD, PhD), P30 AG079280 (PI Jessica Langbaum, PhD), P30 AG062422 (PI Gil Rabinovici, MD), P30 AG066511 (PI Allan Levey, MD, PhD), P30 AG072946 (PI Linda Van Eldik, PhD), P30 AG062715 (PI Sanjay Asthana, MD, FRCP), P30 AG072973 (PI Russell Swerdlow, MD), P30 AG066506 (PI Glenn Smith, PhD, ABPP), P30 AG066508 (PI Stephen Strittmatter, MD, PhD), P30 AG066515 (PI Victor Henderson, MD, MS), P30 AG072947 (PI Suzanne Craft, PhD), P30 AG072931 (PI Henry Paulson, MD, PhD), P30 AG066546 (PI Sudha Seshadri, MD), P30 AG086401 (PI Erik Roberson, MD, PhD), P30 AG086404 (PI Gary Rosenberg, MD), P20 AG068082 (PI Angela Jefferson, PhD), P30 AG072958 (PI Heather Whitson, MD), P30 AG072959 (PI James Leverenz, MD).

Some authors’ affiliation (KS, YN, TI), “*Dementia Inclusion and Therapeutics*,” is an endowed department at the University of Tokyo Hospital funded by Effissimo Capital Management Pte Ltd.

AI-assisted tools were used for language editing; authors take full responsibility for the content.

## Consent Statement

N/A.

## Data Availability

The NACC study data is available from website (https://www.naccdata.org) upon reasonable request.

