## supplementary material for "MMSE-CDR-SB residual as an exploratory indicator of deviation from an Alzheimer’s disease-typical cognitive-functional pattern"

### Supplementary Materials

#### Supplementary Methods

*Detailed cohort construction*

The NACC UDS was treated as a visit-based dataset for cohort construction. For the neuropathological cohort, we first identified deceased participants with available autopsy data. For each participant, we selected the clinical visit closest to death and retained only visits with a non-negative gap between visit date and death date and an interval of 24 months or less. This restriction was chosen to maximize temporal proximity between ante-mortem clinical assessment and post-mortem neuropathological findings. This initial selection yielded 4,412 participants before MMSE restriction.

For the clinical cohort, we identified participants with a clinician-reported primary etiologic diagnosis (*NACCETPR*) and selected the visit closest to death without imposing an upper limit on the interval to death. This approach was chosen to preserve sample size while still capturing a late-disease clinical phenotype. This initial selection yielded 6,414 unique participants before MMSE restriction.

The main analyses were then restricted to visits with MMSE scores between 16 and 30, following the observation by Brogaard et al. that the MMSE-CDR-SB mapping performs less well in more severe stages [Brogaard2025]. After this restriction, the final analytic samples were 1,981 participants in the neuropathological cohort and 3,184 participants in the clinical cohort. Cognitively normal individuals within this MMSE range were retained to preserve the cognitive-functional spectrum. Figure S1 shows the sample selection process.

*Detailed definition of the discrepancy index*

The discrepancy index was calculated at each eligible visit as: *residual = observed CDR-SB − (20.2 − 0.68 × MMSE)*, using the AD-oriented reference equation proposed by Brogaard et al. [Brogaard2025]. A positive residual indicates that observed CDR-SB is higher than expected for a given MMSE value. A negative residual indicates that observed CDR-SB is lower than expected.

For descriptive and stratified analyses, residual values were categorized a priori into 4 bins: ≤−4, −4 to 0, 0 to +4, and ≥+4. These cutoffs were chosen pragmatically for interpretability. Values around 0 indicate concordance with the AD reference line. Negative and positive values indicate deviation below and above the reference line, respectively. Cutoffs at ±4 were used to distinguish modest from more marked departures while preserving adequate sample size within each MMSE-band × residual-bin stratum.

MMSE was also categorized into 3 bands (16–21, 22–27, and 28–30) for descriptive and stratified analyses. These MMSE bands were used in both cross-sectional analyses.

*Detailed coding of neuropathological variables*

Neuropathological variables were derived from NACC autopsy form items using prespecified definitions. Amyloid pathology (A+) was defined as positive if at least 1 of the following criteria was met: Thal phase ≥3, CERAD neuritic plaque score ≥2, or AD neuropathological change (ADNC) score ≥2 [Montine2012]. AD-type tau pathology (T+) was defined as Braak stage ≥3 [Braak1991]. Non-AD tau pathology was defined from PSP, CBD, or ARTAG-related tau variables [Kovacs2016]. Neurodegeneration (N+) was defined as the presence of cortical atrophy, hippocampal atrophy, or hippocampal sclerosis on neuropathological examination [Jack2018].

Additional binary flags were created for Lewy body pathology [McKeith2017], TDP-43 pathology [Nelson2019], hippocampal sclerosis [Nelson2011], and cerebral amyloid angiopathy [vanVeluw2020] from the corresponding NACC neuropathology variables. High vascular burden was defined as a study-specific composite across 6 vascular lesion variables, with a score ≥2 indicating high burden [Skrobot2016]. The source variables, thresholds, and coding rules for all neuropathological variables are listed in Table S1.

*Detailed coding of clinical variables*

Clinical diagnoses were derived from the clinician-reported primary etiologic diagnosis variable (*NACCETPR*). This variable was converted into one-vs-rest binary flags for AD, LBD, VaD, FTLD-other, PSP, CBD, other neurological disorders, and cognitively normal status. These diagnosis flags were based on NACC-recorded etiologic diagnoses and were not re-adjudicated in the present study. Detailed mapping rules are provided in Table S1.

*Descriptive trajectory plots*

For descriptive visualization, we plotted subgroup-specific smoothed trajectories of CDR-SB against MMSE, with the Brogaard AD reference line overlaid. For binary pathological markers and binary clinical diagnosis indicators, trajectories were estimated using linear models with B-spline terms for MMSE (3 degrees of freedom), with CDR-SB as the dependent variable. These plots were used only for descriptive purposes and not for formal inference.

*Cross-sectional descriptive prevalence analyses*

For each binary outcome, visits were cross-classified into MMSE band × discrepancy-bin cells. Crude prevalence was calculated directly within each cell. Adjusted prevalence was estimated using logistic-regression standardization (g-computation). For each outcome, the fitted model had the following form: $logit\{P(Y = 1)\} = \beta_0 + \beta_1(MMSE\ Band) + \beta_2(Residual\ Bin) + \beta_3(Age\ at\ visit) + \beta_4(Sex) + \beta_5(Years\ of\ education)$.

Predicted probabilities were then averaged over the observed covariate distribution within each MMSE band to obtain covariate-adjusted prevalence estimates for each discrepancy bin. If the adjusted model did not converge or analyzable sample size was too small ($n < 20$), crude proportions were reported instead. For adjusted prevalence estimates, 95% confidence intervals were obtained by bootstrap resampling at the participant level (*NACCID*) with 200 replicates.

*Cross-sectional regression analyses*

To quantify associations between the discrepancy index and each neuropathological or clinical outcome, we fitted separate multivariable logistic regression models. The general model form was: $logit\{P(Y = 1)\} = \beta_0 + \beta_1(MMSE\ Band) + \beta_2(Residual\ Bin) + \beta_3(Age\ at\ visit) + \beta_4(Sex) + \beta_5(Years\ of\ education)$.

Models were fitted separately for each pathological flag and each clinical diagnosis flag. No MMSE-band × discrepancy-bin interaction term was included in the main cross-sectional models. In both cohorts, the reference discrepancy category was 0 to +4, and the reference MMSE band was 16–21. Odds ratios, 95% confidence intervals, and p values were derived using cluster-robust standard errors clustered by participant (*NACCID*). Overall likelihood ratio tests were used to assess the contribution of MMSE band and discrepancy bin. Table 2 reports the discrepancy-bin coefficients only, whereas Table S2 provides the full coefficient tables and model summaries.

*Sensitivity analysis for death/autopsy-related selection bias*

To address potential selection bias related to death and autopsy availability, we used the full clinical dataset to examine whether death status was associated with clinician-assigned diagnoses and whether this association varied across discrepancy bins. For each clinical diagnosis outcome, we fitted a logistic regression model of the form: $logit\{P(Y = 1)\} = \gamma_0 + \gamma_1(Residual\ Bin) + \gamma_2(MMSE\ Band) + \gamma_3(Death\ status) + \gamma_4(Age\ at\ visit) + \gamma_5(Sex) + \gamma_6(Years\ of\ education) + \gamma_7[Death\ status \times Residual\ Bin]$.

Odds ratios and Wald p values for the main effect of death status were derived using cluster-robust standard errors clustered by participant. The death status × residual-bin interaction was evaluated using a nested likelihood ratio test. These results are reported in Table S3.

*Exploratory discrimination analyses*

Exploratory discrimination analyses based on the residual alone were also performed. These included MMSE-specific area under the curve (AUC) analyses and smoothed bootstrap confidence bands. Because these analyses were exploratory and not central to the main study aims, they are reported only in the Supplementary Results.

**Table S1: Definition of neuropathological and clinical variables used in the study:**

| Flag / Variable | Source NACC Variable(s) | Positive if: | Definition |
|---|---|---|---|
| **PATHOLOGY FLAGS** | | | |
| ***ATN Framework*** | | | |
| Amyloid positive (A+) | *NPTHAL, NACCNEUR, NPADNC* | Thal phase ≥ 3 OR<br>CERAD score >= 2 OR ADNC >= 2 | Indicates the presence of amyloid-beta plaques in the brain, the hallmark protein aggregate of AD |
| AD-tau positive (T+) | *NACCBRAA* | Braak stage ≥ 3 (stages 3-6) | Reflects neurofibrillary tangle burden following the AD-type tau propagation pattern (Braak & Braak staging) |
| Non-AD tau positive | Derived non-AD tau flag (from *NACCPROG, NACCCBD, NPARTAG*) | PSP tau = 1 OR CBD tau = 1 OR ARTAG = 1 | Captures tau pathology that does not follow the AD Braak pattern, including 4R-tauopathies (PSP, CBD) and age-related tau astrogliopathy (ARTAG). |
| Tau classification (T_class) | *NACCBRAA, NACCPROG, NACCCBD, NPARTAG* | Factor: tau- / AD-tau+ / nonAD-tau+ / unknown | Four-level categorical variable classifying each subject's predominant tau pathology type, enabling distinction between AD-type and non-AD-type tauopathies in comparative analyses. |
| Neurodegeneration (N+) | *NPGRCCA, NPGRHA, NPHIPSCL* | Cortical atrophy ≥ 1 OR Hippocampal atrophy ≥ 1 OR Hippocampal sclerosis in {1,2,3} | Indexes structural neurodegeneration through gross cortical or hippocampal atrophy on neuropathological examination, or the presence of hippocampal sclerosis as a marker of severe neuronal loss. |
| ***Co-pathology Flags*** | | | |
| Lewy body pathology | *NACCLEWY* | *NACCLEWY* = non-zero (i.e., 1-5) | Characterized by abnormal intraneuronal aggregates of alpha-synuclein (Lewy bodies and neurites), ranging from brainstem-predominant to diffuse neocortical distribution; commonly co-occurs with AD pathology. |
| TDP-43 pathology (FTLD-like TDP) | *NPFTDTDP* | *NPFTDTDP* = non-zero (i.e.,1) | TAR DNA-binding protein 43 pathology derived from the NACC overall TDP-43 indicator. In this dataset, this variable predominantly reflects FTLD-type TDP-43 pathology rather than age-related (LATE-type) TDP-43, given that it is based on a single composite item (*NPFTDTDP*) without regional specificity. It should not be interpreted as a broad age-related TDP-43 measure. |
| LATE-like (binary) | NPTDPB, NPTDPC, NPTDPD, NPTDPE, NPALSMND | Regional TDP-43 present in amygdala (*NPTDPB*=1), hippocampus (*NPTDPC*=1), or entorhinal/inferior temporal cortex (*NPTDPD*=1); excluding cases with FTLD-type TDP or ALS/MND (*NPALSMND*=1) | Binary indicator of LATE-like TDP-43 pathology, defined using regional TDP-43 variables following a limbic-predominant distribution consistent with LATE neuropathological change. Cases with concurrent FTLD-type TDP-43 or ALS/motor neuron disease were excluded to minimize FTLD contamination. |

| Flag / Variable | Source NACC Variable(s) | Positive if: | Definition |
|---|---|---|---|
| LATE stage ≥2 (binary) | NPTDPB, NPTDPC, NPTDPD, NPTDPE, NPALSMND | LATE_stage3 ≥ 2, i.e., amygdala (*NPTDPB*=1) AND hippocampus (*NPTDPC*=1) affected; excluding FTLD/ALS cases | Binary indicator derived from a 0–3 ordinal staging scheme approximating the LATE staging framework: stage 0 = no regional TDP-43; stage 1 = amygdala only; stage 2 = amygdala + hippocampus; stage 3 = amygdala + hippocampus + neocortex (*NPTDPE*=1). Stage ≥2 was used as the threshold for logistic models, reflecting involvement beyond the amygdala. Cases with FTLD-type TDP-43 or ALS/MND were excluded. |
| Hippocampal sclerosis (HS) | *NPHIPSCL* | *NPHIPSCL* = non-zero (i.e., 1-3) | HS is defined by severe neuronal loss and gliosis in the CA1/subiculum region of the hippocampus; in older adults it is associated with TDP-43 pathology (LATE) and presents clinically with amnestic dementia mimicking AD. |
| Cerebral amyloid angiopathy (CAA) | *NACCAMY* | *NACCAMY* = non-zero (i.e., 1-3) | CAA refers to the deposition of amyloid-beta in the walls of cortical and leptomeningeal vessels, predisposing to lobar hemorrhages and white matter injury; it frequently co-exists with parenchymal AD pathology. |
| High vascular burden (VascHigh) | *NACCARTE, NACCMICR, NACCINF, NACCHEM, NPOLDD, NPWMR* | *VascScore* ≥ 2 (sum of 6 binary vascular features) | Composite indicator of significant cerebrovascular pathology, aggregating arteriolosclerosis, microinfarcts, large infarcts, hemorrhages, old macroscopic infarcts, and white matter rarefaction; a score >= 2 denotes a high burden likely to contribute to cognitive impairment. |
| **CLINICAL (ETIOLOGIC DIAGNOSIS) FLAGS** | | | |
| ***Clinical Diagnosis Mapping*** | | | |
| Clinical diagnosis code label (NACCETPR_dx) | *NACCETPR* | Integer codes 1-30, 88, 90 -> string labels (e.g., 1 -> '01_AD') | Human-readable label derived from the NACC etiologic primary diagnosis code. |
| Binary diagnosis flag (dx_{diagnosis}) | | 1 if NACCETPR_dx == target; 0 otherwise; NA if NACCETPR_dx is NA | One-vs-rest binary indicator for a specific clinical diagnosis. |
| **VISIT SELECTION (NEAR-DEATH)** | **Constructed from / Selection rule** | | |
| Neuropathological cohort visit selection (*d_near_pathol*) | *VISITYR, VISITMO, VISITDAY, NACCDIED, NACCYOD, NACCMOD,* autopsy data variables | nearest visit before death (gap_days ≥ 0), restricted to gap_months ≤ 24 | Among deceased participants with autopsy data, selects the single clinical visit closest to death and within 24 months before death for neuropathological analyses. |

| Flag / Variable | Source NACC Variable(s) | Positive if: | Definition |
|---|---|---|---|
| Clinical cohort visit selection (*d_near_clin*) | *VISITYR, VISITMO, VISITDAY, NACCDIED, NACCYOD, NACCMOD, NACCETPR* | nearest visit before death (gap_days ≥ 0), with no upper limit on gap_months | Among participants with a clinician-reported primary etiologic diagnosis, selects the single clinical visit closest to death without a time restriction for clinical diagnosis analyses. |

Summary of neuropathological and clinical variables derived from the National Alzheimer's Coordinating Center (NACC) dataset. For each variable or flag used in the analyses, the table lists the corresponding source NACC variable(s), the criterion used to define positivity (coded as 1), and a brief description of the construct. LATE-like variables were available only for participants assessed with neuropathological form version 10 or 11 (*NPFORMVER* = 10 or 11; n = 511 with complete data). Participants with *NPFORMVER* = NA (n = 1,470) had no regional TDP-43 data and were therefore coded as NA for all LATE-related variables. This substantial missingness should be considered when interpreting LATE-related results.

**Abbreviations:** AD, Alzheimer's disease; ADNC, Alzheimer's disease neuropathologic change; ALS, amyotrophic lateral sclerosis; ARTAG, aging-related tau astrogliopathy; Aβ, amyloid-beta; ATN, amyloid/tau/neurodegeneration framework; CAA, cerebral amyloid angiopathy; CBD, corticobasal degeneration; CERAD, Consortium to Establish a Registry for Alzheimer's Disease; FTLD, frontotemporal lobar degeneration; FTLD-TDP, frontotemporal lobar degeneration with TDP-43 pathology; HS, hippocampal sclerosis; LATE, limbic-predominant age-related TDP-43 encephalopathy; LBD, Lewy body disease; NACC, National Alzheimer's Coordinating Center; NACCID, National Alzheimer's Coordinating Center participant identifier; PSP, progressive supranuclear palsy; TDP-43, TAR DNA-binding protein 43.

##### Table S2: Complete Results of GLM Analysis (Table 2)

#### A. Neuropathological cohort

| Term | A+ (amyloid) | T+ (AD tau) | Non-AD tau+ | N+ (neurodegeneration) | TDP-43 pathology | Hippocampal sclerosis | Lewy body pathology | High vascular burden |
|---|---|---|---|---|---|---|---|---|
| *N (n positive)* | *1023 (553)* | *1010 (635)* | *1018 (31)* | *511 (396)* | *428 (18)* | *507 (58)* | *1020 (229)* | *1023 (475)* |
| **MMSE 22–27 vs 16–21 (ref)** | **0.61 (0.43–0.87) p = 0.006** | 0.71 (0.47–1.06) p = 0.092 | 1.02 (0.40–2.62) p = 0.961 | 0.78 (0.37–1.62) p = 0.507 | 1.15 (0.36–3.64) p = 0.811 | 0.74 (0.37–1.46) p = 0.380 | 0.77 (0.53–1.12) p = 0.166 | **1.53 (1.08–2.16) p = 0.016** |
| **MMSE 28–30 vs 16–21 (ref)** | **0.37 (0.26–0.53) p = <0.001** | **0.28 (0.19–0.41) p = <0.001** | 1.21 (0.49–2.99) p = 0.687 | **0.33 (0.17–0.64) p = 0.001** | **0.04 (0.01–0.30) p = 0.002** | **0.16 (0.07–0.39) p = <0.001** | **0.51 (0.34–0.76) p = <0.001** | 1.22 (0.87–1.72) p = 0.251 |
| Age at visit (per year) | **1.03 (1.02–1.05) p = <0.001** | **1.06 (1.05–1.08) p = <0.001** | **0.95 (0.92–0.98) p = <0.001** | **1.05 (1.03–1.08) p = <0.001** | **0.87 (0.82–0.94) p = <0.001** | 1.02 (0.98–1.06) p = 0.278 | 1.01 (1.00–1.02) p = 0.193 | **1.04 (1.03–1.06) p = <0.001** |
| Sex: female vs male (ref) | 0.88 (0.68–1.14) p = 0.321 | 1.08 (0.82–1.43) p = 0.578 | 0.62 (0.28–1.37) p = 0.237 | 1.03 (0.65–1.64) p = 0.884 | 0.96 (0.26–3.46) p = 0.947 | 0.77 (0.42–1.40) p = 0.395 | **0.57 (0.41–0.77) p = <0.001** | 1.10 (0.85–1.43) p = 0.468 |
| Education (per year) | 0.99 (0.97–1.01) p = 0.545 | 0.98 (0.96–1.01) p = 0.226 | 0.97 (0.92–1.03) p = 0.309 | 0.95 (0.88–1.03) p = 0.231 | 1.00 (0.84–1.19) p = 0.968 | 0.98 (0.87–1.09) p = 0.667 | 0.99 (0.97–1.02) p = 0.634 | 1.01 (0.99–1.04) p = 0.275 |

#### B. Clinical cohort

| Term | AD | LBD (clinical) | VaD | FTLD-other | PSP | CBD | Cognitively normal |
|---|---|---|---|---|---|---|---|
| *N (n positive)* | *3072 (1525)* | *3072 (119)* | *3072 (109)* | *3072 (161)* | *3072 (38)* | *3072 (16)* | *3072 (935)* |
| **MMSE 22–27 vs 16–21 (ref)** | **0.36 (0.29–0.45) p = <0.001** | **0.50 (0.33–0.78) p = 0.002** | 1.51 (0.90–2.55) p = 0.121 | 0.99 (0.64–1.52) p = 0.951 | **2.99 (1.16–7.71) p = 0.024** | 1.01 (0.31–3.28) p = 0.992 | **14.81 (9.38–23.38) p = <0.001** |

| Term | AD | LBD (clinical) | VaD | FTLD-other | PSP | CBD | Cognitively normal |
|---|---|---|---|---|---|---|---|
| **MMSE 28–30 vs 16–21 (ref)** | **0.04 (0.03–0.06) p = <0.001** | **0.23 (0.13–0.41) p = <0.001** | 1.19 (0.67–2.12) p = 0.560 | **0.36 (0.21–0.63) p = <0.001** | 1.38 (0.46–4.13) p = 0.565 | 0.49 (0.13–1.93) p = 0.311 | **292.75 (183.69–466.55) p = <0.001** |
| Age at visit (per year) | **1.04 (1.03–1.05) p = <0.001** | **0.98 (0.97–1.00) p = 0.015** | **1.05 (1.03–1.08) p = <0.001** | **0.88 (0.86–0.89) p = <0.001** | **0.92 (0.91–0.94) p = <0.001** | **0.92 (0.89–0.94) p = <0.001** | **1.02 (1.01–1.03) p = <0.001** |
| Sex: female vs male (ref) | 0.96 (0.81–1.14) p = 0.644 | **0.24 (0.15–0.39) p = <0.001** | 0.76 (0.52–1.12) p = 0.164 | 0.67 (0.44–1.01) p = 0.058 | 0.98 (0.49–1.97) p = 0.965 | 1.25 (0.47–3.34) p = 0.659 | **1.53 (1.23–1.91) p = <0.001** |
| Education (per year) | 0.99 (0.98–1.00) p = 0.146 | 1.00 (0.97–1.03) p = 0.955 | 0.95 (0.90–1.01) p = 0.119 | **1.02 (1.01–1.04) p = 0.010** | 1.02 (1.00–1.04) p = 0.061 | 1.02 (0.99–1.04) p = 0.228 | 0.98 (0.96–1.00) p = 0.097 |

The complete results of the logistic regression models reported in Table 2. While Table 2 reports odds ratios for the CDR–MMSE discrepancy bins only, this table provides the OR (95% CI) and p-value for all remaining model terms: MMSE band (reference: 16–21) and covariates (age at visit, sex, and years of education). Each cell displays the odds ratio followed by the 95% confidence interval in parentheses and the associated p-value. Models were adjusted for all listed variables simultaneously, with cluster-robust standard errors clustered by participant (*NACCID*). N (n positive) indicates the number of observations with complete data and the number of outcome-positive observations respectively. Bold values indicate statistical significance ($p < 0.05$). NC* = not calculable due to extreme separation.

**Part A** presents results from the neuropathological cohort (n = 1,981), restricted to participants with autopsy data and a clinical visit within 24 months prior to death. **Part B** presents results from the clinical cohort (n = 3,184), including participants with a clinician-reported etiologic diagnosis at the visit closest to death.

**Abbreviations:** MMSE, Mini-Mental State Examination; ref, reference category; A+, amyloid-positive; T+, Alzheimer's disease tau-positive; Non-AD tau+, non-Alzheimer tau-positive; N+, neurodegeneration-positive; TDP-43, TAR DNA-binding protein 43; HS, hippocampal sclerosis; LBD, Lewy body disease; VascHigh, high vascular burden; AD, Alzheimer's disease; VaD, vascular dementia; FTLD, frontotemporal lobar degeneration; FTLD-other, other frontotemporal lobar degeneration; PSP, progressive supranuclear palsy; CBD, corticobasal degeneration.

#### Table S3: Sensitivity Analysis: Mortality Bias

##### A. Mortality bias test: main effect and interaction with residual bin

| Diagnosis | N | p (died main effect) | p (died × bin interaction, LRT) |
|---|---|---|---|
| AD | 20,430 | **<0.001** | **<0.001** |
| Lewy body disease (clinical) | 20,430 | **0.044** | 0.707 |
| Vascular dementia | 20,430 | 0.247 | **0.010** |
| FTLD-other | 20,430 | **0.004** | **0.003** |
| PSP | 20,430 | 0.547 | 0.258 |
| CBD | 20,430 | 0.274 | 0.661 |
| Other neurological | 20,430 | **<0.001** | 0.331 |
| Cognitively normal | 20,430 | **<0.001** | **<0.001** |

##### B. Odds ratio for death status (died = 1 vs. 0) per diagnosis

| Diagnosis | OR (died vs. alive) | 95% CI | z | p-value |
|---|---|---|---|---|
| AD | 0.35 | 0.23–0.52 | -5.07 | **<0.001** |
| CBD | 3.01 | 0.42–21.69 | 1.09 | 0.274 |
| FTLD-other | 3.07 | 1.44–6.54 | 2.90 | **0.004** |
| Lewy body disease (clinical) | 3.32 | 1.03–10.65 | 2.02 | **0.044** |
| Normal cognition | 5.82 | 2.90–11.68 | 4.96 | **<0.001** |
| Other neurological | 0.00 | 0.00–0.00 | -49.43 | **<0.001** |
| PSP | 1.71 | 0.30–9.72 | 0.60 | 0.547 |
| Vascular dementia | 0.59 | 0.24–1.44 | -1.16 | 0.247 |

Sensitivity analyses evaluating potential mortality-related selection bias in the association between the MMSE–CDR-SB discrepancy index and clinical diagnoses. Analyses were conducted in the full clinical dataset using logistic regression models including discrepancy bin, MMSE band, death status (died = 1/0), age, sex, and years of education.

**Table S3A** reports p-values testing the association between death status and each clinical diagnosis outcome, as well as the interaction between death status and discrepancy bin. The column *p (died main effect)* corresponds to the Wald test for the main effect of death status. The column *p (died × bin interaction)* corresponds to a likelihood ratio test (LRT) comparing models with and without the interaction between death status and discrepancy bin.

**Table S3B** reports the odds ratio (OR) for death status (died vs. alive) for each clinical diagnosis derived from the same logistic regression models. OR values greater than 1 indicate higher odds of the diagnosis among deceased participants, whereas values lower than 1 indicate lower odds.

N represents the complete-case sample size available for each diagnosis outcome. Cluster-robust standard errors were used with clustering at the participant level (*NACCID*). Confidence intervals are reported on the odds ratio scale.

**Abbreviations:** OR, odds ratio; CI, confidence interval; LRT, likelihood ratio test; MMSE, Mini-Mental State Examination; CDR-SB, Clinical Dementia Rating Sum of Boxes; AD, Alzheimer's disease; FTLD, frontotemporal lobar degeneration; PSP, progressive supranuclear palsy; CBD, corticobasal degeneration.

**Table S4: Supplementary interaction analyses: AD background x residual bin**

**A. Summary of interaction tests (AD background x residual bin, LRT)**

| Outcome | N | Positive | p (co_AD main effect) | p (co_AD x resid_bin, LRT) |
|---|---|---|---|---|
| TDP (FTLD-like) | 422 | 18 | 0.043 | 0.064 |
| LATE_like | 361 | 96 | 0.637 | 0.774 |
| LATE_stage_ge2 | 341 | 57 | 0.971 | 0.336 |
| VascHigh | 499 | 303 | 0.866 | 0.889 |

**B.** OR for AD background (co_AD) main effect

| Outcome | OR | 95% CI | p |
|---|---|---|---|
| TDP (FTLD-like) | 0.11 | 0.01-0.94 | 0.043 |
| LATE_like | 0.67 | 0.13-3.46 | 0.637 |
| LATE_stage_ge2 | 0.97 | 0.19-5.02 | 0.971 |
| VascHigh | 0.90 | 0.27-3.00 | 0.866 |

**C.** GLM associations between residual bin and LATE variables (adjusted)

| Outcome | Resid bin | OR | 95% CI | p |
|---|---|---|---|---|
| LATE_like | -4 to 0 | 2.62 | 0.32-21.6 | 0.372 |
| LATE_like | 0 to 4 | 4.27 | 0.52-34.8 | 0.175 |
| LATE_like | ≥ + 4 | 4.53 | 0.52-39.8 | 0.173 |
| LATE_stage_ge2 | -4 to 0 | 1.33 | 0.17-10.8 | 0.786 |
| LATE_stage_ge2 | 0 to 4 | 3.19 | 0.41-25.0 | 0.270 |
| LATE_stage_ge2 | ≥ + 4 | 2.90 | 0.34-24.6 | 0.329 |

Reference category: resid_bin ≤ -4. Models adjusted for MMSE band, age, sex, education. Cluster-robust SE clustered by NACCID.

Table S4 presents supplementary interaction analyses examining whether the association between the residual index and selected neuropathological variables differ by AD neuropathological background (co_AD, defined as ADNC ≥ 2). Analyses were conducted in the neuropathological cohort using logistic regression models adjusted for MMSE band, age at visit, sex and years of education, with cluster-robust standard errors clustered by participants (NACCID).

Table S4A reports the sample size, number of positive cases, p -value for the main effect of co_AD, and likelihood ratio test (LRT) p-value for the co_AD x residual bin interaction term.

Table S4B reports the odds ratio (OR) and 95% confidence interval for the main effect of co_AD (co_AD = 1 vs 0) from the interaction model.

Table S4C reports adjusted ORs for residual bin categories relative to the reference bin (≤−4) for LATE-like and LATE stage ≥2, derived from models without the interaction term. Wide confidence intervals reflect small case numbers and substantial missingness in LATE-related variables, which were available only for participants assessed with neuropathological form version 10 or 11 (*NPFORMVER* = 10–11; n = 511).

**Abbreviations:** OR, odds ratio; CI, confidence interval; LRT, likelihood ratio test; ADNC, Alzheimer's disease neuropathological change; MMSE, Mini-Mental State Examination; NACCID, National Alzheimer's Coordinating Center participant identifier; TDP, TAR DNA-binding protein 43; LATE, limbic-predominant age-related TDP-43 encephalopathy; VascHigh, high vascular burden.

**Table S5: Longitudinal association between pathological variables and residual trajectory**

| Outcome | Term | β | 95% CI | p |
|---|---|---|---|---|
| A+ (amyloid) | t_yrs (baseline slope) | 0.073 | 0.039–0.107 | <0.001 |
| A+ (amyloid) | t_yrs × Apos_AD | 0.072 | 0.031–0.112 | 0.001 |
| TDP-43 (FTLD-like) | t_yrs (baseline slope) | 0.086 | 0.064–0.107 | <0.001 |
| TDP-43 (FTLD-like) | t_yrs × TDP | 0.142 | 0.032–0.253 | 0.012 |
| LATE-like | t_yrs (baseline slope) | 0.065 | 0.038–0.093 | <0.001 |
| LATE-like | t_yrs × LATE_like | 0.086 | 0.042–0.130 | <0.001 |
| LATE stage ≥2 | t_yrs (baseline slope) | 0.073 | 0.047–0.099 | <0.001 |
| LATE stage ≥2 | t_yrs × LATE_stage_ge2 | 0.087 | 0.036–0.138 | 0.001 |
| VascHigh | t_yrs (baseline slope) | 0.123 | 0.092–0.154 | <0.001 |
| VascHigh | t_yrs × VascHigh | −0.004 | −0.043–0.034 | 0.822 |

β = regression coefficient from linear mixed-effects model (lme4::lmer, ML). t_yrs = years from first visit. Reference group for interaction: pathology absent. All models adjusted for age at visit, sex, and years of education, with random intercept and slope by participant (NACCID).

Results of exploratory linear mixed-effects models examining whether the longitudinal trajectory of the residual index differed by neuropathological group. Each model included a time × pathology interaction term, adjusted for age at visit, sex, and years of education, with random intercepts and slopes by participant (*NACCID*). Only the time slope and interaction terms are shown.

**Abbreviations:** β, regression coefficient; CI, confidence interval; t_yrs, years from first eligible visit; TDP, TDP-43 pathology (FTLD-like); LATE, limbic-predominant age-related TDP-43 encephalopathy; VascHigh, high vascular burden; NACCID, National Alzheimer's Coordinating Center participant identifier.

**Figure S1: Study sample selection flow chart**

Flowchart illustrating the selection of participants from the National Alzheimer's Coordinating Center (NACC) Uniform Data Set (UDS) between 2005 and 2023. Participants were excluded if they had a known autosomal dominant Alzheimer's disease mutation, history of stroke, Parkinson's disease, alcohol abuse, schizophrenia, depressive disorder of less than two years' duration, or bipolar disorder. Eligible participants were then allocated to two analytic cohorts according to the availability of data. The neuropathological cohort included participants with available autopsy data and a clinical visit within 24 months prior to death, restricted to MMSE scores between 16 and 30 (n = 1,981). The clinical cohort included participants with a clinician-reported etiologic diagnosis and MMSE scores between 16 and 30 at the visit closest to death (n = 3,184).

**Abbreviations:** NACC, National Alzheimer's Coordinating Center; UDS, Uniform Data Set; MMSE, Mini-Mental State Examination.

**Figure S2: Number of positive cases across CDR-SB/MMSE discrepancy bins, stratified by MMSE band in the neuropathological cohort**

Bar plots display the number of participants positive for each neuropathological variable across categories of discrepancy between observed and expected CDR-SB given MMSE (discrepancy bins: ≤−4, −4 to 0, 0 to +4, ≥+4), stratified by MMSE band (16–21, 22–27, 28–30). Only variables with n ≥ 10 cases and used in our GLM analysis are shown. Raw counts are displayed above each bar. A+, amyloid-positive (Thal phase ≥ 3 or CERAD score >= 2 OR ADNC >= 2); T+, AD-tau positive (Braak stage ≥ 3); Non-AD tau+, tau-positive in a non-AD distribution; N+, neurodegeneration-positive.

**Abbreviations:** CDR-SB, Clinical Dementia Rating Scale Sum of Boxes; MMSE, Mini-Mental State Examination.

**Figure S3: Number of positive cases across CDR-SB/MMSE discrepancy bins, stratified by MMSE band in the clinical cohort**

Bar plots display the number of participants positive for each clinician-reported etiologic diagnosis across categories of discrepancy between observed and expected CDR-SB given MMSE (discrepancy bins: ≤−4, −4 to 0, 0 to +4, ≥+4), stratified by MMSE band (16–21, 22–27, 28–30). Only variables with n ≥ 10 cases overall are shown. Raw counts are displayed above each bar.

**Abbreviations:** CDR-SB, Clinical Dementia Rating Scale Sum of Boxes; MMSE, Mini-Mental State Examination; AD, Alzheimer's disease; LBD, Lewy body dementia (clinical diagnosis); VaD, vascular dementia; FTLD-other, frontotemporal lobar degeneration (other subtypes); PSP, progressive supranuclear palsy; CBD, corticobasal degeneration; Cogn. normal, cognitively normal.

**Figure S4: Prevalence of selected pathologies across residual bins, stratified by AD background**

Bar plots display the crude proportion of participants positive for each pathological variable (FTLD-like TDP, LATE-like, LATE stage ≥ 2, high vascular burden) across residual bins (≤−4, −4 to 0, 0 to +4, ≥+4), separately for participants with (co_AD=1) and without (co_AD=0) high AD neuropathological change (ADNC ≥ 2). Sample sizes (n) are shown above each bar. Crude rather than adjusted proportions are shown due to sparse data in AD-stratified subgroups. Analyses were restricted to the neuropathological cohort. These figures correspond to the supplementary analysis reported in Table S4.

**Figure S5: Longitudinal trajectories of the MMSE-CDR-SB residual by pathological group**

Longitudinal trajectories of the MMSE–CDR-SB residual index in participants with and without each neuropathological marker. Trajectories were estimated from linear mixed-effects models adjusted for age, sex, and education, with random intercepts and slopes by participant. Shaded areas represent 95% confidence intervals. *VascHigh* is not shown (interaction p = 0.822). Analyses included participants with ≥2 eligible visits (neuropathological cohort: 3,642 participants, 14,598 visits). LATE-related variables available only for NPFORMVER 10–11.

**Abbreviations:** CDR-SB, Clinical Dementia Rating Sum of Boxes; MMSE, Mini-Mental State Examination; TDP-43, TAR DNA-binding protein 43 (FTLD-like variable); LATE, limbic-predominant age-related TDP-43 encephalopathy; NPFORMVER, neuropathological form version.
